# Junctional Hounsfield unit ratio: understanding patient-specific vertebral bone strength for proximal junctional kyphosis risk assessment in adult spinal deformity surgery

**DOI:** 10.64898/2026.04.05.26349586

**Authors:** Yasuhiro Nagatani, Naoki Segi, Sadayuki Ito, Jun Ouchida, Ippei Yamauchi, Yukihito Ode, Yuya Okada, Yosuke Takeichi, Hiroto Tachi, Yujiro Kagami, Kazuaki Morishita, Ryotaro Oishi, Yuichi Miyairi, Yoshinori Morita, Kazuma Ohshima, Hiroki Oyama, Keisuke Ogura, Ryuichi Shinjo, Tetsuya Ohara, Taichi Tsuji, Tokumi Kanemura, Shiro Imagama, Hiroaki Nakashima

## Abstract

**Study design:** A retrospective case control study

**Objective:** To predict proximal junctional kyphosis (PJK) risk by normalizing individual vertebral bone strength using the ratio of vertebral Hounsfield unit (HU) values around the upper instrumented vertebrae (UIV).

**Summary of background data:** PJK poses a significant challenge in treating patients after adult spinal deformity (ASD) surgery. While the vertebral body HU value is associated with PJK risk, the optimal threshold remains unclear, and a relative assessment of HU values within individuals has not been conducted.

**Methods:** Data on patients who underwent corrective fusion of the middle to lower thoracic region of the pelvis for ASD were assessed. The 126 patients were categorized into PJK and non-PJK groups. We compared the patients’ backgrounds, vertebral body HU, and junctional HU ratio, defined as the HU value of UIV+1 divided by the HU value of UIV (HU_UIV+1_/HU_UIV_). The UIV+2/UIV+1 HU ratio was calculated similarly.

**Results:** The PJK and non-PJK groups included 30 and 96 patients, respectively. After propensity score matching, 28 patients from each group were analyzed. HU values at UIV+2 and UIV+1 (117.0 ± 46.6 vs 145.1 ± 45.9, p=0.018, and 105.5 ± 36.2 vs 147.3 ± 44.9, p<0.001, respectively) were lower in the PJK group. Junctional HU ratio was significantly lower in the PJK group (0.88 ± 0.18 vs 1.13 ± 0.25, p<0.001), and receiver operating characteristic analysis showed that the junctional HU ratio had the highest discriminative ability (area under the curve 0.812). At the optimal cutoff value (HU ratio of 0.905), the sensitivity and specificity for PJK were 64.3% and 89.3%, respectively.

**Conclusions:** A low junctional HU ratio was strongly associated with PJK after ASD surgery. This parameter reflects the bone strength mismatch at the proximal junction and may help improve preoperative risk assessment and UIV selection.

## Introduction

Adult spinal deformity (ASD) surgery, particularly when it involves long-segment fusion, is becoming more common owing to the increasing awareness of how spinal deformities impact quality of life^1^. Despite advances in surgical techniques and perioperative patient management, these procedures can still lead to postoperative complications. One significant complication is proximal junctional kyphosis (PJK), which occurs near the upper end of a spinal fusion construct^2^, causing pain and functional impairment, and frequently necessitating revision surgery^3^. PJK is particularly concerning because of its high incidence and severe clinical outcomes, with some studies reporting rates as high as 40%^4^. Consequently, PJK poses a significant challenge in managing patients after ASD surgery.

The causes of PJK are diverse; however, one of the primary risk factors is low bone mineral density (BMD)^5^. Methods for assessing BMD include dual-energy X-ray absorptiometry (DEXA) and computed tomography (CT) values (Hounsfield units, HU)^6^. DEXA has traditionally been considered the gold standard for assessing BMD. However, spinal degeneration and vascular calcification can lead to elevated T-score obtained from DEXA, making it challenging to accurately measure spinal bone strength^7, 8^. However, measuring bone density using HU values enables the selection of specific vertebrae within the CT scan and is gaining attention as a new way to assess spinal bone strength^9–12^. Furthermore, recent studies have shown that low HU values at critical vertebral levels, particularly in the upper instrumented vertebra (UIV) and adjacent vertebrae (UIV+1), are strongly associated with PJK^13^. These findings indicate that bone strength around the UIV is crucial for preventing PJK.

However, as HU values differ with individual bone density and vertebral level^14^, the optimal HU threshold for predicting PJK remains unclear. Importantly, PJK may not simply result from low bone strength alone; rather, it results from a mismatch in bone strength between the rigid fused segment and adjacent mobile segments. Therefore, evaluating the relative difference in bone strength across the junction, instead of relying solely on absolute HU values, may offer a more clinically relevant means of selecting optimal UIV and preventing PJK. Here, we hypothesized that PJK can occur because of a mechanical gap between the fused construct and the mobile spine. Thus, we propose that assessing the relative strength within the patient’s individual spinal continuum is necessary to understand this mechanical gap, rather than simply evaluating the HU values of each vertebra. Therefore, the objective of this study was to clarify the relationship between vertebral HU values around the UIV and their gradations, specifically, the “junctional HU ratio” and the occurrence of PJK in ASD surgery, thereby improving the preoperative risk assessment for patients undergoing ASD.

## Material and Methods

### Study Population

In this retrospective multicenter study, we reviewed the data of patients who underwent corrective spinal surgery for ASD between February 2009 and February 2020. The inclusion criteria were: (1) aged ≥ 18 years, (2) patients who met at least one of the following radiographic criteria (Cobb angle > 20°, sagittal vertical axis [SVA] > 5 cm, pelvic tilt [PT] > 25°, or thoracic kyphosis [TK] > 60°); (3) patients who received corrective fusion from the middle to lower thoracic to the pelvis, and (4) minimum of 2-year follow-up period. The exclusion criteria were: (1) fresh vertebral fracture, (2) presence of infection, and (3) presence of a tumor. Patients with insufficient data were excluded. Among the 214 patients in the initial cohort, 126 were included in the final analysis (Figure 1).

**Figure 1.**
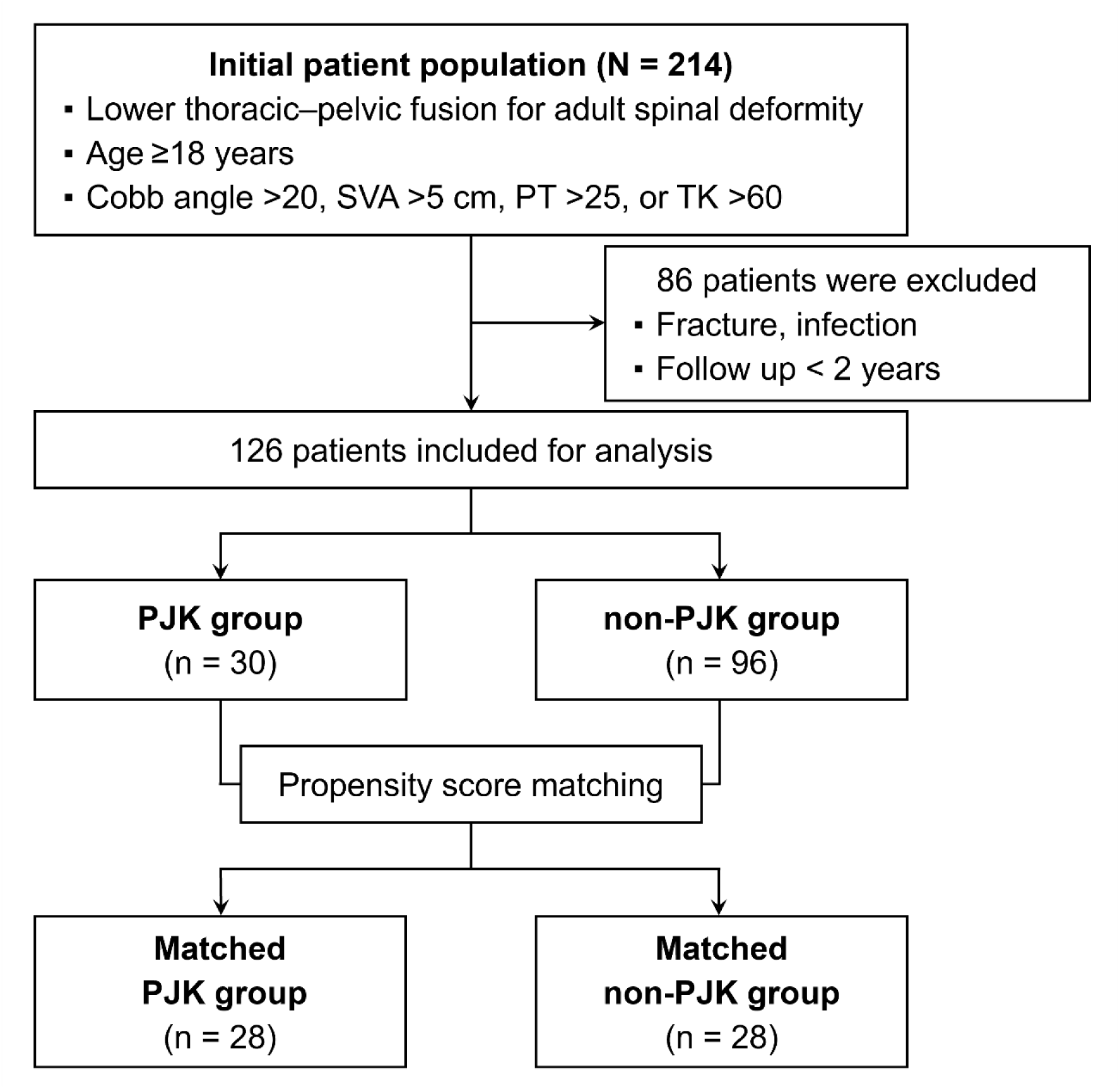
Patient selection flowchart. SVA, sagittal vertical axis; PT, pelvic tilt; TK, thoracic kyphosis; PJK, proximal junctional kyphosis.

This study was approved by the institutions affiliated with the authors. All procedures followed the principles of the Declaration of Helsinki. The requirement for informed consent was waived due to the study’s retrospective nature.

### Variables and Grouping

Age, sex, body mass index (BMI), Japanese Orthopaedic Association (JOA) score^15^, spinal alignment parameters assessed using spine full-length standing X-rays, such as SVA, TK, lumbar lordosis (LL), PT, sacral slope (SS), pelvic incidence (PI), and the types of PJK were investigated. Full-length standing spine X-ray images were obtained in a freestanding position, with the fingers resting on the clavicles while looking straight ahead.

CT scans were used to assess bone quality, and HU values were measured at the UIV, UIV+1, UIV+2, and L4 vertebrae. For instance, the HU value at the UIV is denoted as HU_UIV_. HU values were calculated using the elliptical tool in the standard PACS software, ensuring that the regions of interest excluded the cortical bone, bone defects, and local lesions. To assess the HU value, the regions of interest were measured on the axial images at three separate locations: immediately inferior to the superior endplate, in the middle of the vertebral body, and superior to the inferior endplate (Figure 2)^16^. Furthermore, the HU value ratios were measured. The junctional HU ratio was defined as the UIV+1/UIV HU ratio, that is, ratio of HU_UIV+1_ to HU_UIV_. The UIV+2/UIV+1 HU value ratio was similarly calculated (Figure 3).

**Figure 2.**
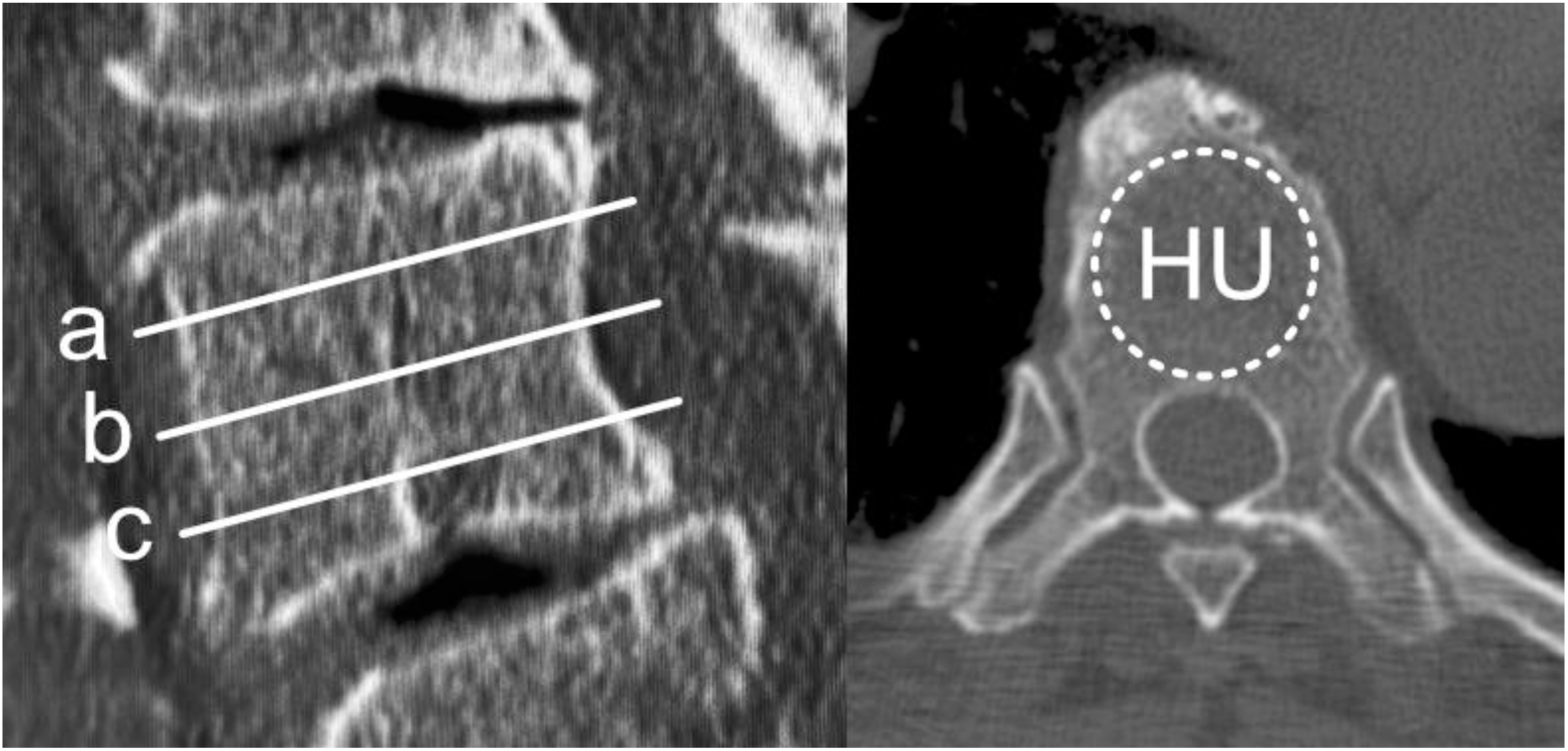
HU measurement method. a. Inferior to the superior end plate b. Middle of the vertebral body c. Superior to the inferior end plate Vertebral Hounsfield unit (HU) value was calculated as the average HU value measured at three locations: a, b, and c. HU, Hounsfield unit.

**Figure 3.**
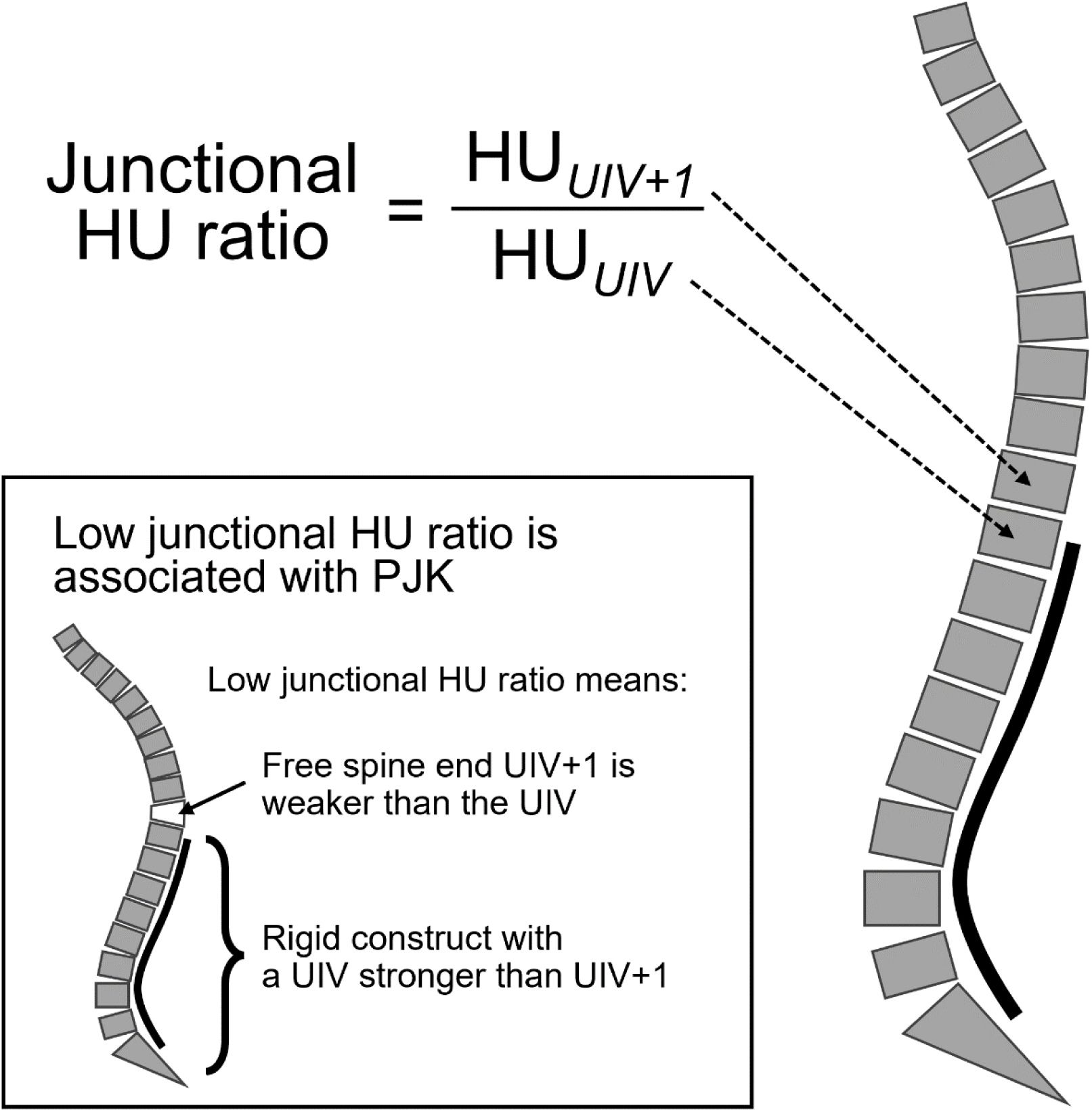
Junctional HU ratio. HU, Hounsfield unit.

Radiographic criteria for PJK were defined as > 10° change from baseline in UIV+2 and > 10° angulation^17^. PJK is divided into three types: type 1, PJK from disc and ligamentous failure; type 2: bone failure; and type 3: implant/bone interface failure^18^. Patients were categorized into two groups: those who developed PJK postoperatively (PJK group, n = 30) and those who did not (non-PJK group, n = 96).

### Statistical Analysis

EZR software was used for statistical analyses (Saitama Medical Center, Jichi Medical University, Japan)^19^. The Mann–Whitney U test was used to compare continuous variables between the PJK and non-PJK groups, while Fisher’s exact test was used for categorical variables. A logistic regression model was used to calculate the odds ratio (OR) with 95% confidence interval (CI) for PJK risk. Propensity score matching was used to balance baseline characteristics across groups. Variables considered for propensity score matching were age, sex, and preoperative SVA. The residuals were checked for normality using a histogram. The Variance Inflation Factor (VIF) metric showed that all adjusted covariates had VIF values < 2.0. Statistical significance was set at p < 0.05. Receiver operating characteristic (ROC) curve analysis was used to investigate the relationship between HU values, the junctional HU ratio, and PJK. The optimal cutoff value was determined using the Youden index.

## Results

We included 126 patients who met the inclusion criteria (Figure 1). The mean age was 70.9 ± 7.3 years, and 104 (82.5%) were women. The mean HU values were 141.2 ± 46.1 for HU_UIV+2_, 138.2 ± 44.6 for HU_UIV+1_, 134.2 ± 44.4 for HU_UIV_, and 156.2 ± 63.6 for HU_L4_. PJK occurred in 30 patients (23.8%), with type 2 PJK being the most common type (n = 13; 43.3%) (Table 1). Some baseline variables, including age and preoperative SVA, were significantly different between the 30 and 96 patients in the PJK and non-PJK groups, respectively (see Tables, Supplemental Digital Contents 1 and 2).

**Table 1.** Patients’ demographics.

|  | **Analytic cohort**<br>**(N = 126)** |
| --- | --- |
| Age (years) | 70.9 ± 7.3 |
| Sex, female | 104 (82.5%) |
| BMI (kg/m<sup>2</sup>) | 23.4 ± 3.9 |
| UIV |  |
| T9 | 11 (8.7%) |
| T10 | 78 (61.9%) |
| T11 | 17 (13.5%) |
| T12 | 20 (15.9%) |
| HU value |  |
| UIV+2 | 141.2 ± 46.1 |
| UIV+1 | 138.2 ± 44.6 |
| UIV | 134.2 ± 44.4 |
| L4 | 156.2 ± 63.6 |
| JOA score |  |
| pre | 15.4 ± 4.9 |
| post 2 years | 22.2 ± 3.9 |
| PJK | 30 (23.8%) |
| PJK type |  |
| Type 1 | 11 (36.7%) |
| Type 2 | 13 (43.3%) |
| Type 3 | 6 (20.0%) |

BMI, body mass index; HU, Hounsfield unit; UIV, upper instrumented vertebrae; JOA, Japanese Orthopaedic Association; PJK, proximal junctional kyphosis.

After propensity score matching, analysis revealed that in the PJK group, mean HU_UIV+2_ (117.0 ± 46.6 vs 145.1 ± 45.9, p = 0.018), HU_UIV+1_ (105.5 ± 36.2 vs 147.3 ± 44.9, p < 0.001) and junctional HU ratio at UIV+1/UIV (0.88 ± 0.18 vs 1.13 ± 0.25, p < 0.001) were significantly lower than that of the non-PJK group. No statistically significant differences were observed between the groups regarding HU_UIV_, preoperative and postoperative 2-year JOA score, or HU ratios at UIV+2/UIV+1 (Table 2). At 2 years postoperatively, the PJK group had significantly greater TK (47.5 ± 12.5 vs 35.4 ± 12.8, p = 0.002) and LL (46.3 ± 10.1 vs 39.6 ± 11.9, p = 0.016). No significant differences were observed in other alignment parameters (Table 3).

**Table 2.** Background characteristics between PJK and non-PJK group after matching.

|  | **PJK group**<br>**(n = 28)** | **non-PJK group**<br>**(n = 28)** | **p value** |
| --- | --- | --- | --- |
| Age (years) | 74.1 ± 4.1 | 74.1 ± 5.0 | 0.86 |
| Sex, female | 24 (85.7%) | 22 (78.6%) | 0.73 |
| BMI (kg/m<sup>2</sup>) | 23.0 ± 3.3 | 23.9 ± 4.0 | 0.77 |
| UIV |  |  | 0.46 |
| T9 | 4 (14.3%) | 3 (10.7%) |  |
| T10 | 20 (71.4%) | 16 (57.1%) |  |
| T11 | 1 (3.6%) | 4 (14.3%) |  |
| T12 | 3 (10.7%) | 5 (17.9%) |  |
| HU |  |  |  |
| UIV+2 | 117.0 ± 46.6 | 145.1 ± 45.9 | **0.018** |
| UIV+1 | 105.5 ± 36.2 | 147.3 ± 44.9 | **< 0.001** |
| UIV | 119.5 ± 32.2 | 137.1 ± 55.1 | 0.30 |
| L4 | 146.8 ± 58.5 | 148.2 ± 53.2 | 0.74 |
| JOA score |  |  |  |
| pre | 13.5 ± 5.0 | 14.4 ± 5.3 | 0.54 |
| post 2 years | 20.8 ± 4.3 | 22.4 ± 3.9 | 0.12 |
| HU ratio |  |  |  |
| UIV+1/UIV | 0.88 ± 0.18 | 1.13 ± 0.25 | **<0.001** |
| UIV+2/UIV+1 | 1.11 ± 0.34 | 0.99 ± 0.13 | 0.50 |

BMI, body mass index; HU, Hounsfield unit; UIV, upper instrumented vertebrae; JOA, Japanese Orthopedic Association; PJK, proximal junctional kyphosis.

**Table 3.** Comparison of spinal alignment between PJK and non-PJK group after matching.

|  |  | **PJK group**<br>**(n = 28)** | **non-PJK group**<br>**(n = 28)** | **p value** |
| --- | --- | --- | --- | --- |
| SVA | Pre | 105.5 ± 41.7 | 106.9 ± 49.4 | 0.99 |
|  | Post 2 years | 59.7 ± 43.0 | 49.0 ± 29.4 | 0.47 |
| TK | Pre | 27.1 ± 17.8 | 22.9 ± 13.3 | 0.46 |
|  | Post 2 years | 47.5 ± 12.5 | 35.4 ± 12.8 | **0.002** |
| LL | Pre | 18.5 ± 16.1 | 15.3 ± 21.7 | 0.65 |
|  | Post 2 years | 46.3 ± 10.1 | 39.6 ± 11.9 | **0.016** |
| PT | Pre | 32.0 ± 9.4 | 31.6 ± 13.1 | 0.99 |
|  | Post 2 years | 23.3 ± 9.8 | 23.4 ± 8.9 | 0.63 |
| SS | Pre | 19.1 ± 10.4 | 21.3 ± 12.8 | 0.50 |
|  | Post 2 years | 28.7 ± 8.9 | 27.3 ± 9.2 | 0.59 |
| PI | Pre | 51.8 ± 10.8 | 52.9 ± 11.9 | 0.61 |
|  | Post 2 years | 52.1 ± 10.3 | 50.7 ± 10.7 | 0.76 |

SVA, sagittal vertical axis; TK, thoracic kyphosis; LL, lumbar lordosis; PT, pelvic tilt; SS, sacral slope; PI, pelvic incidence; PJK, proximal junctional kyphosis.

Multivariable analysis showed that the junctional HU ratio at UIV+1/UIV was associated with the PJK group, with an OR of 0.00202 (95% CI: 0.0000298–0.138, p = 0.004). The HU ratio of UIV+2/UIV+1 was not associated with PJK (Table 4). ROC analysis showed that the junctional HU ratio had the highest discriminative ability (area under the curve [AUC] 0.812) than HU_UIV_ (AUC 0.582), HU_UIV+1_ (AUC 0.787), and HU_UIV+2_ (AUC 0.689). At the optimal cutoff value (HU ratio, 0.905), the sensitivity and specificity were 64.3% and 89.3%, respectively (Figure 4).

**Figure 4.**
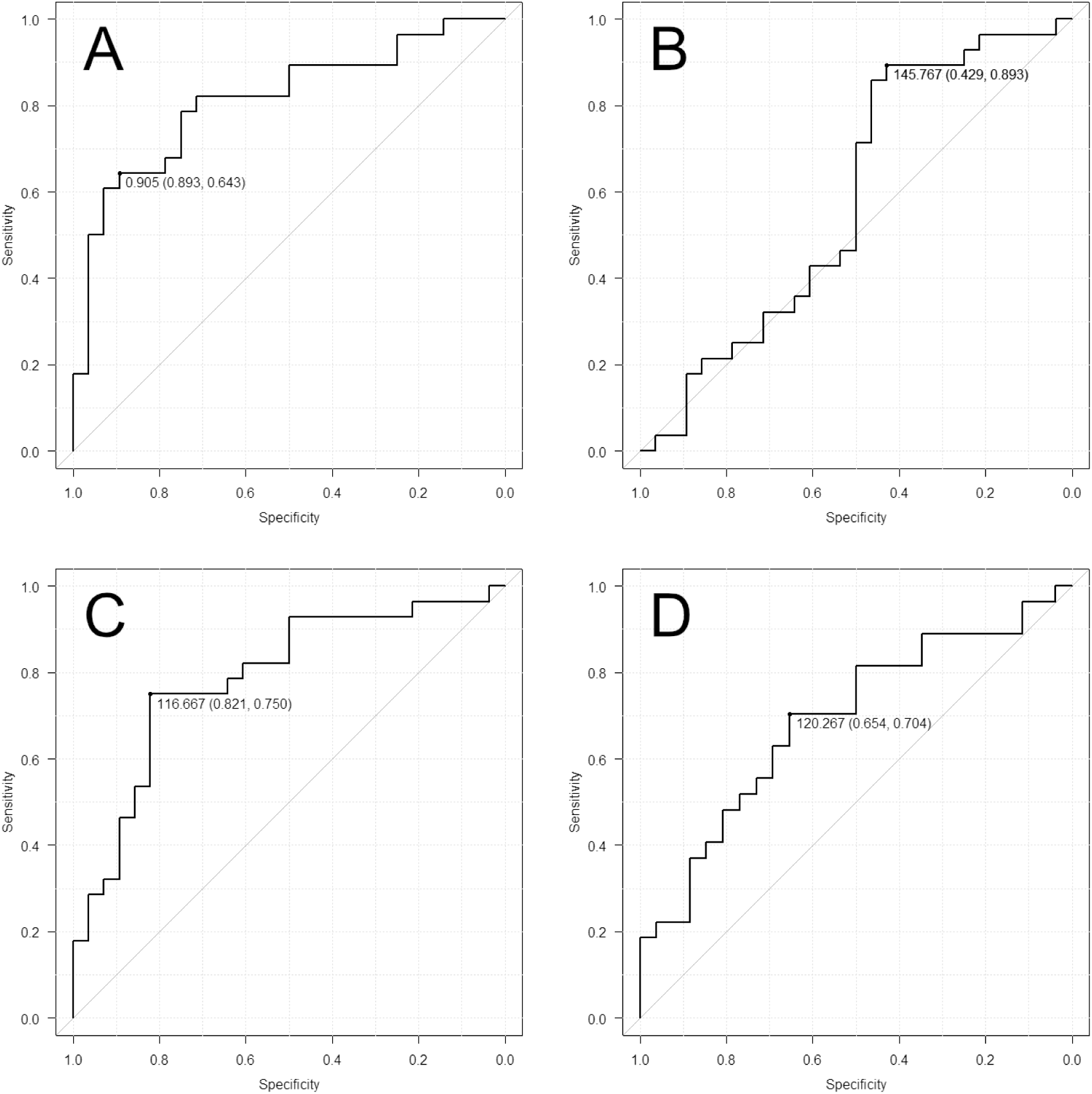
Receiver operating characteristic curves for vertebral Hounsfield unit parameters in proximal junctional kyphosis. A. Junctional Hounsfield unit (HU) ratio (HU_UIV+1_/HU_UIV_) showed the highest discriminative ability (area under the curve (AUC) = 0.812). B. HU_UIV_ (AUC = 0.582). C. HU_UIV+1_ (AUC = 0.787). D. HU_UIV+2_ (AUC = 0.689). HU, Hounsfield unit.

**Table 4.** Multivariable linear regression analysis for PJK after matching.

| **Variables** | **OR [95% CI]** | **p value** |
| --- | --- | --- |
| Age | 0.975 [0.843 – 1.130] | 0.73 |
| Sex, female | 1.330 [0.240 – 7.350] | 0.75 |
| BMI | 0.965 [0.798 – 1.170] | 0.72 |
| Junctional HU ratio (UIV+1/UIV) | 0.00202 [0.0000298 – 0.138] | **0.004** |
| HU ratio (UIV+2/UIV+1) | 3.000 [0.161 – 55.800] | 0.46 |

BMI, body mass index; HU, Hounsfield unit; UIV, upper instrumented vertebrae; PJK, proximal junctional kyphosis; SVA, sagittal vertical axis; CI, confidence interval.

## Discussion

This multicenter study showed that a lower junctional HU ratio, defined as the ratio of the HU value at UIV+1 to that at UIV, was independently associated with PJK development after ASD surgery. Importantly, this parameter showed superior discriminative ability compared to absolute HU values alone, indicating that the relative mismatch in bone strength across the proximal junction is crucial in PJK development.

Previous studies have consistently reported that low vertebral HU values at or near the UIV are associated with an increased risk of PJK^13, 20^; however, absolute HU values do not fully capture the structural transition that occurs between the rigid fused segment and the adjacent mobile spine. This study builds on previous findings by showing that the relative difference in bone strength between adjacent vertebrae, rather than absolute bone density alone, is independently associated with PJK. This finding supports the concept that mechanical complications (MCs) at the proximal junction may arise from a structural mismatch, where the adjacent cranial vertebra lacks sufficient strength to handle the increased mechanical demands imposed by the rigid fusion construct. Osteoporosis increases PJK risk^13^, vertebral HU values correlate with osteoporosis assessment conducted via DEXA, and low vertebral HU values are associated with PJK risk^30, 31^. Furthermore, PJK can occur because of a mechanical gap between the fused construct and the mobile spine^21^. We concluded that a comprehensive understanding of this mechanical gap requires evaluating the relative strength within the patient’s spinal continuum, rather than independently assessing and comparing the HU values of each vertebra. Consequently, we devised a junctional HU ratio that normalizes individual bone strength.

The findings of this study offer a new perspective on selecting UIV. While vertebral HU values generally increase from the lower to the upper thoracic spine, patients with osteoporosis demonstrated a decrease in HU values from T9 to T7^14, 22^. Previous reports indicate that setting the upper thoracic spine as the UIV reduces PJK risk^23, 24^. Our findings indicate that setting the UIV above T7–9 (where bone strength is reduced) may help reduce PJK risk. T9 or T10 is often selected as the UIV; however, the selection criteria remain unclear^25^. Thus, the junctional HU ratio may be a practical tool for surgical planning and risk stratification. Surgeons may consider modifying the surgical strategy when the ratio is low, especially when the junctional HU ratio is < 0.9. Potential approaches include selecting a cranial UIV with more favorable bone strength, using prophylactic vertebral augmentation, reinforcing the posterior ligamentous structures, or adjusting the magnitude of alignment correction. These individualized strategies may help lessen mechanical mismatches and reduce PJK risk. Importantly, since the HU values can be easily obtained from routine preoperative CT scans, this parameter can be incorporated into clinical practice without incurring extra costs or exposing patients to additional radiation.

Notably, our findings must also be interpreted in the context of sagittal alignment correction. Even after propensity score matching, the PJK group showed significantly greater postoperative thoracic kyphosis and lumbar lordosis than the non-PJK group did. These differences imply that the alignment correction itself may have contributed to the increased mechanical stress at the proximal junction. Previous studies have shown that excessive correction, particularly when not in harmony with the patient’s intrinsic spinal alignment, increases the risk of MCs^26–29^. Therefore, PJK development is likely to be multifactorial and involves bone strength mismatch and alignment-related mechanical stresses. Thus, the junctional HU ratio should be considered part of a comprehensive risk assessment that includes global alignment parameters.

The relationship between the junctional HU ratio and the different types of PJK warrants further consideration. Reports examining the relationship between HU values and PJK type are limited; nevertheless, the results of this study imply that the junctional HU ratio may be involved in the occurrence of multiple PJK types. PJK is classified into three morphological types^18^: type 1, disc and ligamentous failure; type 2, bone failure; and type 3, implant-bone interface failure. In this study, type 2 PJK, primarily due to bone failure, was the most common, indicating an influence of low HU values around the UIV. Incidence rates of PJK subtypes vary, with some studies indicating that type 1 is more common, while others show that type 2 predominates^18, 32^. However, even among the type 1 cases showing disc and ligamentous failure, some CT cases demonstrated mild vertebral deformation at UIV+1. Therefore, cases classified as type 1 may include instances that cannot be explained solely by pure disc or ligamentous failure. Furthermore, bone density at UIV+1 may decrease postoperatively compared to preoperatively, and reduced HU values in adjacent vertebrae can be risk factors for vertebral fractures and disc degeneration^33, 34^. Therefore, in cases with a low junctional HU ratio, there is a possibility that the ratio may decrease further after fixation surgery. This decline could induce minor vertebral deformations at UIV+1, potentially developing type 1 PJK. In this study, the combined frequency of types 1 and 2 PJK was 80.0% (24 and 30 cases, respectively), showing that the junctional HU ratio may be significant in the occurrence of type 1 and type 2 PJK.

This study had some limitations. First, its retrospective design limited the ability to establish causality. Second, propensity score matching was conducted; nonetheless, residual confounding factors may remain. Particularly, sagittal alignment parameters were not included in the multivariable model because of concerns about the sample size and multicollinearity. Future studies with larger cohorts should incorporate the bone strength and alignment variables to clarify their relative contributions. Additionally, in multivariable analysis, while HU_UIV+1_, HU_UIV+2_, and junctional HU ratio were significantly associated with PJK in univariate analyses, it is not feasible to include these metrics simultaneously in a multivariable model because of multicollinearity (variance inflation factor > 20). Third, bone mineral density measured using DEXA was not available for all patients, preventing a direct comparison between HU values and conventional osteoporosis assessments. Finally, the surgical techniques, implant types, and postoperative management were not fully standardized across the participating institutions, which may have affected the outcomes. Despite these limitations, this study offers valuable insights into the biomechanical and clinical significance of bone strength mismatch at the proximal junction. The junctional HU ratio is a novel patient-specific parameter that reflects the structural transition between the fused and mobile spinal segments. When interpreted alongside sagittal alignment correction, this parameter may provide helpful guidance for surgical planning and risk stratification.

## Conclusions

In conclusion, the junctional HU ratio is a novel patient-specific parameter that reflects the relative bone strength across the proximal junction. A low junctional HU ratio, especially one below approximately 0.9, may indicate an increased PJK risk. Assessing this ratio using preoperative CT may help guide optimal UIV selection and reduce the risk of mechanical complications after ASD surgery.

## Data Availability

The datasets generated and/or analyzed in the current study are available from the corresponding author upon reasonable request.

## Ethical Approval

The study protocol was approved by the Human Research Ethics Committee and Institutional Review Board of Nagoya University (No.2016-0177). This study was performed in accordance with the principles of the Declaration of Helsinki.

## Acknowledgments

We thank Editage (www.editage.jp) for English editing. The authors disclose the use of ChatGPT (OpenAI) during the manuscript preparation. This tool was used for drafting assistance, including the identification of potential methodological limitations and missing analyses. All the outputs were carefully evaluated, verified, and edited by the authors. All authors are responsible for the accuracy and integrity of this study.

---

**Supplemental Table 1.** Background characteristics between PJK and non-PJK group.

|  | **PJK group**<br>**(n = 30)** | **non-PJK group**<br>**(n = 96)** | **p value** |
| --- | --- | --- | --- |
| Age (years) | 74.3 ± 4.1 | 69.9 ± 7.7 | **0.003** |
| Sex, female | 26 (86.7%) | 78 (81.3%) | 0.59 |
| BMI (kg/m<sup>2</sup>) | 23.0 ± 3.4 | 23.5 ± 4.1 | 0.92 |
| UIV |  |  | 0.46 |
| T9 | 4 (13.3%) | 7 (7.3%) |  |
| T10 | 20 (66.7%) | 58 (60.4%) |  |
| T11 | 2 (6.7%) | 15 (15.6%) |  |
| T12 | 4 (13.3%) | 16 (16.7%) |  |
| HU |  |  |  |
| UIV+2 | 116.1 ± 45.1 | 149.5 ± 43.6 | **< 0.001** |
| UIV+1 | 106.7 ± 35.3 | 148.1 ± 42.8 | **< 0.001** |
| UIV | 118.6 ± 31.3 | 139.3 ± 46.9 | **0.031** |
| L4 | 149.7 ± 59.3 | 158.0 ± 65.1 | 0.42 |
| JOA score |  |  |  |
| pre | 15.1 ± 5.9 | 15.4 ± 4.7 | 0.88 |
| post 2 years | 20.9 ± 4.3 | 22.6 ± 3.8 | **0.042** |
| HU ratio |  |  |  |
| UIV+1/UIV | 0.90 ± 0.19 | 1.11 ± 0.24 | **< 0.001** |
| UIV+2/UIV+1 | 1.09 ± 0.34 | 1.01 ± 0.20 | 0.67 |

BMI, body mass index; HU, Hounsfield unit; UIV, upper instrumented vertebrae; JOA, Japanese Orthopedic Association; PJK, proximal junctional kyphosis.

**Supplemental Table 2.** Comparison of spinal alignment between PJK and non-PJK group.

|  |  | **PJK group**<br>**(n = 30)** | **non-PJK group**<br>**(n = 96)** | **p value** |
| --- | --- | --- | --- | --- |
| SVA | Pre | 111.1 ± 45.6 | 85.8 ± 54.3 | **0.027** |
|  | Post 2 years | 62.9 ± 43.3 | 44.0 ± 42.3 | **0.038** |
| TK | Pre | 26.0 ± 18.2 | 21.6 ± 14.8 | 0.23 |
|  | Post 2 years | 46.4 ± 14.0 | 34.0 ± 12.1 | **<0.001** |
| LL | Pre | 17.1 ± 16.7 | 17.1 ± 19.4 | 0.98 |
|  | Post 2 years | 45.1 ± 11.3 | 40.1 ± 12.4 | **0.010** |
| PT | Pre | 32.3 ± 9.2 | 30.9 ± 12.1 | 0.55 |
|  | Post 2 years | 23.3 ± 9.5 | 22.7 ± 7.8 | 0.48 |
| SS | Pre | 19.0 ± 10.1 | 20.0 ± 12.0 | 0.85 |
|  | Post 2 years | 28.7 ± 8.6 | 28.0 ± 9.7 | 0.67 |
| PI | Pre | 51.9 ± 10.4 | 50.8 ± 12.1 | 0.55 |
|  | Post 2 years | 52.0 ± 9.9 | 50.6 ± 11.5 | 0.68 |

SVA, sagittal vertical axis; TK, thoracic kyphosis; LL, lumbar lordosis; PT, pelvic tilt; SS, sacral slope; PI, pelvic incidence; PJK, proximal junctional kyphosis.

